# A Multiplex qPCR to Differentiate Monkeypox Virus Clades

**DOI:** 10.1101/2025.02.07.25321854

**Authors:** Christopher T. Williams, Alessandra Romero-Ramirez, Adeleye Adesola Semiu, Samuel Oluwafunmbi Ifabumuyi, Susan Gould, Dominic Wooding, Collette Allen, Anushri Somasundaran, Nicodemus Nnabuike Mkpuma, Dorcas Gado, Jolly Amoche Adole, Abdulakeem Eniola Amoo, Abisola Ajoke Adeyemi, Laure Bosquillon de Jarcy, Christine Goffinet, Jake Dunning, Malcolm G Semple, ISARIC CCP UK investigators, Esto Bahizire, Afolabi Akinpelu, Thomas E Fletcher, Ana Cubas-Atienzar, Cristina Leggio, Adeyinka Adedeji, Adesuyi A. Omoare, Thomas Edwards

**Affiliations:** Centre for Drugs and Diagnostics, Liverpool School of Tropical Medicine, Liverpool, United Kingdom; National Reference Laboratory Department of Public Health Laboratory Services (PHLS), Nigeria Centre for Disease Control and Prevention (NCDC), Gaduwa, Abuja 900110, Nigeria; UK Health Security Agency, Porton Down–Salisbury, UK; Department of Infectious and Transboundary animal diseases, National Veterinary Research Institute, Vom, Plateau State, Nigeria; Virology Division, National Veterinary Research Institute, Vom, Plateau State, Nigeria; Central Public Health Laboratory, Yaba Lagos, Nigeria Centre for Disease Control and Prevention (NCDC); Tropical Disease Biology, Liverpool School of Tropical Medicine, Liverpool, United Kingdom; Pandemic Sciences Institute, University of Oxford, UK; Infectious Diseases Department, Royal Free London NHS Foundation Trust, London, UK; Institute of Infection, Veterinary and Ecological Sciences, University of Liverpool, Liverpool, UK; The International Severe Acute Respiratory and emerging Infection Consortium (ISARIC); Catholic University of Bukavu - Bugabo Campus, Mission Ave, Bukavu, Kinshasa, Democratic Republic of Congo; Clinical Sciences, Liverpool School of Tropical Medicine, Liverpool, United Kingdom

## Abstract

We designed a multiplex qPCR to differentiate monkeypox virus clades. In evaluations using clinical samples collected in the UK and Nigeria, the assay had an overall sensitivity of 78% (95% CI: 67.67% to 86.14%) and specificity of 94% (95% CI: 80.84% to 99.30%); for samples under Ct35 sensitivity was 98% (95% CI: 91.72% to 99.96%) and specificity was 94% (95% CI: 80.84% to 99.30%).

## Introduction

Mpox is a viral zoonosis caused by the monkeypox virus (MPXV). There are two mpox clades with clade I typically associated with a higher disease severity and case fatality ratio compared with clade II [1].

Clade II is subdivided into IIa and IIb. Lineage B.1, a newly emerged lineage of IIb, was responsible for the 2022 global outbreak. Lineage B.1 is more transmissible than previously observed, potentially driven by human adaptation [2], with significant human-to-human transmission reported, particularly in men who have sex with men (MSM)[3, 4]. Lower mortality rates were recorded during the 2022 outbreak, potentially driven by the population demographics and health systems involved [5]. Clade I is subdivided into Ia and Ib, with Ib emerging during an outbreak in the Democratic Republic of the Congo (DRC) in 2023. Much like Lineage B.1, clade Ib infection causes lower mortality than previous clade I outbreaks whilst being more infectious, with sustained human-to-human transmission [6] linked to sexual contact [7]. A ∼1kb deletion occurs in the OPG032 gene in clade Ib [6], where the US CDC clade II PCR assay is located.

Mpox diagnosis is typically by PCR performed on swabs taken from lesion scabs or respiratory samples [8]. As mpox presents with similar clinical features between clades, clade identification typically requires sequencing which can be time-consuming, expensive and difficult to implement in low- and middle-income countries (LMICs). Clade/lineage identification is important due to differences in disease severity and for epidemiological purposes, particularly with new outbreak strains.

### The study

For an initial evaluation, we used DNA extracted from cultured Lineage B.1 MPXV (Slovenia_MPXV-1_2022 strain, Clade hMPXV-1, lineage B.1, European Virus Archive), as well as 47 clinical samples from the ISARIC Clinical Characterisation Protocol study (ethical approval; National Research Ethics Service and Health Research Authority IRAS ID:126600, REC 13/SC/0149). Thirty-two samples were MPXV-positive by the US CDC reference PCR [9] and 15 were negative.

Eleven samples were collected from participants with West Africa travel-associated mpox in 2018 and 36 from UK participants infected with Lineage B.1 mpox in 2022. Samples were classified by clade using the geographic source and date of collection; all confirmed UK cases from 2018 were from West Africa and infected with clade II mpox, whilst UKHSA sequencing data revealed that 99% of mpox samples from the 2022 cohort were Lineage B.1. [10] DNA was extracted using the QIAamp 96 Virus QIAcube HT Kit (Qiagen, UK) on a QIAcube HT (Qiagen, UK) following manufacturer’s instructions.

We also tested 54 MPXV-positive samples collected as part of routine mpox diagnostic surveillance at the National Reference Laboratory (NRL) of the Nigerian Centre for Disease Control and Prevention (NCDC) Abuja, Nigeria (approval was granted by the Research Governance Unit at NCDC). Samples were collected in 2017 (n=7), 2018 (n=11), 2019 (n=10), 2021 (n=10), 2023 (n=12) and 2024 (n=4). Twenty Varicella Zoster Virus (VZV) qPCR-positive samples were used as negatives. DNA was extracted using a QIAmp DNA mini kit (Qiagen, UK) following manufacturer’s instructions.

Clade-specific SNPs were identified using Nextstrain (https://nextstrain.org/mpox/all-clades). Sequences for target genes for each clade/lineage were downloaded from NCBI Genbank and aligned by ClustalW using MEGA 11. Probes were manually designed to contain the SNP(s) in the middles of the probe. Primers were designed using PrimerQuest (IDT DNA). The Clade I probe targets the F3L gene (T46417C, G46421A, A46427C, C46435T), and the lineage B.1 probe targets OPG109 (C84587T). We designed an assay on the OPG210 gene (G183695A, C183696T) to distinguish IIb from ‘non-IIb’; non-IIb includes IIa and Clade I which contain wild-type nucleotides where IIb contains two mutations. An assay targeting the ∼1 kb deletion (Δ19,128–20,270) in the OPG032 gene of clade Ib was designed after assay evaluations. See Table 1 for results interpretation.

**Table 1.** Assay interpretation

| Result | Probe |  |  |  |  |
| --- | --- | --- | --- | --- | --- |
|  | Clade I | Clade Ib | Non-IIb | Clade IIb | Lineage B.1 |
| Clade I | Positive | Negative | Positive | Negative | Negative |
| Clade Ib | Positive | Positive | Positive | Negative | Negative |
| Clade IIa | Negative | Negative | Positive | Negative | Negative |
| Clade IIb | Negative | Negative | Negative | Positive | Negative |
| Lineage B.1 | Negative | Negative | Negative | Positive | Positive |

**Table 2.**
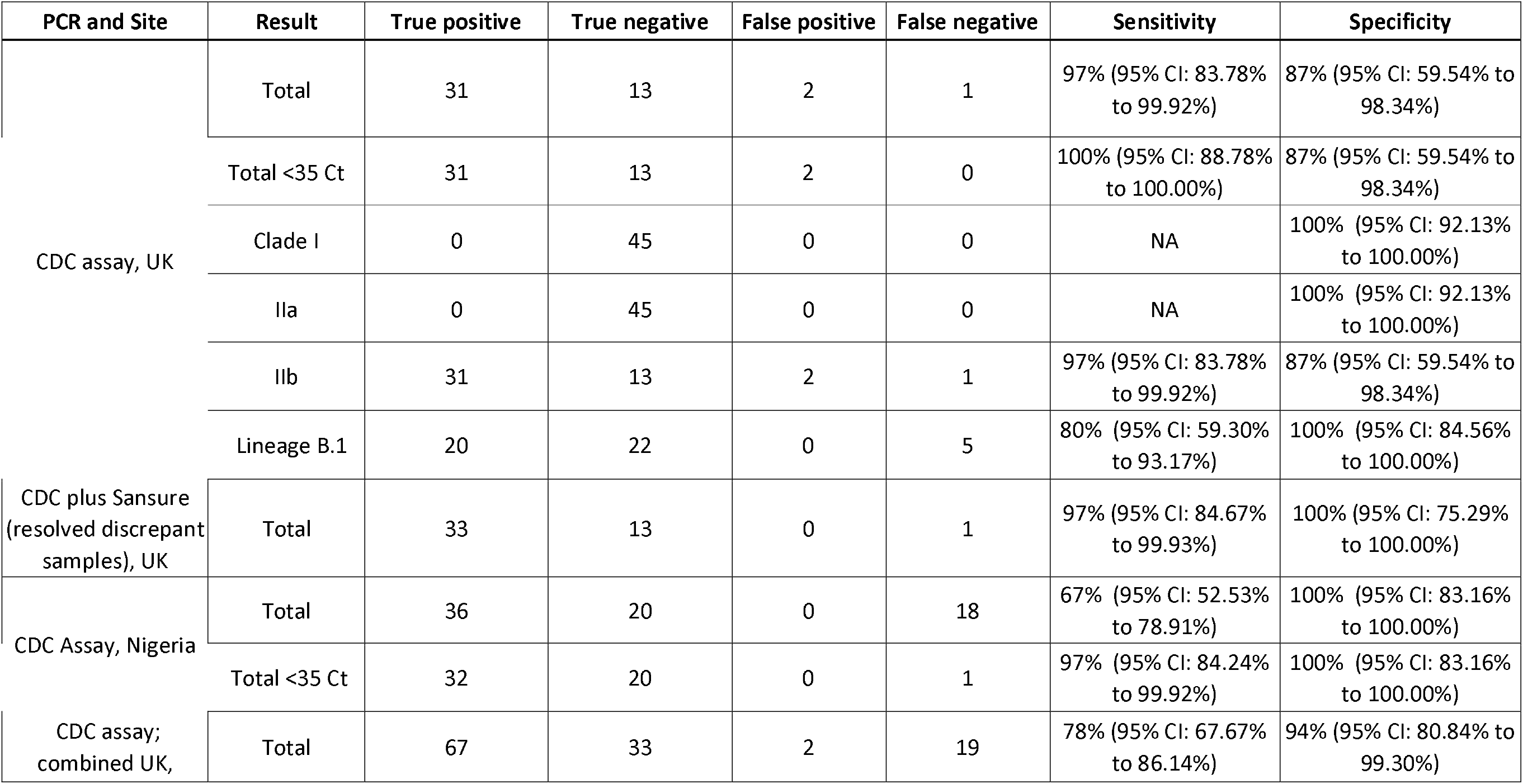
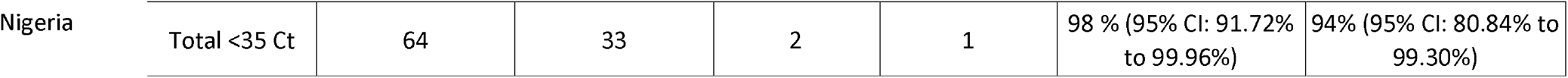
Evaluation results

qPCR assays were performed in a 12.5 μl reaction volume, containing TaqPath Fast mix (Applied Biosystems, US), differing concentrations of primers and probes (Supplementary Table 1), nuclease-free H2O and 2.5 μl DNA. To improve specificity for the Lineage B.1 assay we designed a ‘blocker’ oligo, which is identical to the B.1 probe, except that it contains the wild-type nucleotide in place of the SNP, and a phosphate at the 5’ end instead of a fluorophore. All experiments were performed on a Quantstudio 5 (Applied Biosystems, US) using the following thermal profile; 95 °C for 20 seconds, followed by 40 cycles of 95 °C for 1 second and 60 °C for 10 seconds. We used a cutoff of Ct 38 for positivity, and thresholds were set at 10% of the maximum fluorescence of the positive control curve.

Standard curves of 500bp synthetic double-stranded DNA containing the amplicons from each target (Twist Bioscience, USA) were performed for clade I, clade Ib, IIb and non-IIb in triplicate from 1×10^7^ copies/μl to 1 copy/μl (Figure 1). DNA was extracted from lineage B.1 culture and quantified into copies/μl using the IIb standard curve to make the standard curve for lineage B1, starting at 1×10^5^ copies/μl to 1 copy/μl. All targets were successfully amplified in three replicates down to 10 copies/μl. Clade I and IIb assays amplified all three replicates for 1 copy/μl; clade IIa, lineage B.1 and clade Ib amplified 2/3 replicates.

**Figure 1.**
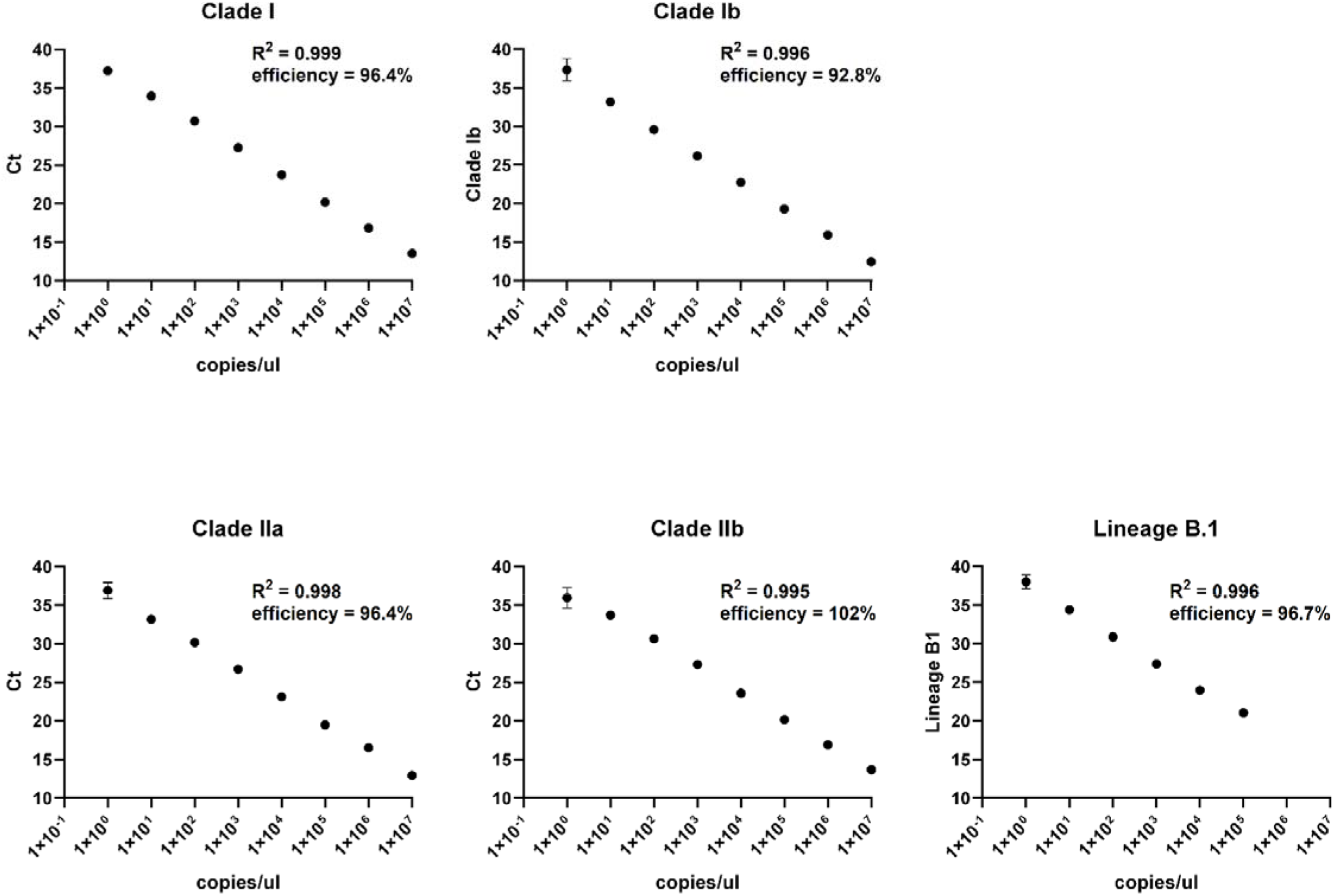
Standard curve of each assay target

For the UK evaluation our assay has 97% (95% CI: 83.78% to 99.92%) sensitivity and 87% (95% CI: 59.54% to 98.34%) specificity compared with the US CDC Mpox qPCR. Discrepant samples were tested using the Sansure Monkeypox virus Nucleic Acid Diagnostic Kit (Sansure Biotech, China) The two false positives and single false negative were positive by Sansure, suggesting two false negative results using the US CDC mpox qPCR. Our assay was 100% specific and 80% sensitive for detecting Lineage B.1. The five false negatives were all Ct34 or higher.

In the evaluation at the NCDC we had an overall sensitivity of 67% (95% CI: 52.53% to 78.91%) and specificity of 100% (95% CI: 83.16% to 100.00%) compared with the US CDC assay. 20% (11/54) of the NCDC samples were >Ct35; typically, Ct values from lesion swabs in the acute phase would be lower [11], with samples >Ct 35 predicted to have no or very little infectivity [12]. When testing samples <Ct35 we observed a sensitivity of 97% (95% CI: 84.24% to 99.92%) and a specificity of 100% (95% CI: 83.16% to 100.00%). All samples were Clade IIb.

## Conclusions

We designed a multiplexed qPCR to identify MPXV clades/lineages in a single reaction and evaluated it on a collection of clade IIb and Lineage B.1 clinical samples, DNA extracted from culture of Lineage B.1, as well as MPXV-negative samples.. Whilst we did not have access to clinical samples containing clade I or clade IIa viruses we are planning further evaluations to address this.

Other PCR assays can distinguish between clade I and clade II. Recently, another qPCR assay to detect clade I, clade II, clade IIb and the outbreak strain B.1 was developed [13], which requires two tubes per sample and does not include clade Ib. Another assay can detect clade Ib in singleplex [14].

The rapid identification of MPXV clades is becoming ever more necessary with the advent of multiple outbreaks attributed to different clades. Whilst sequencing remains the gold standard, PCR-based methods remain a useful tool when access to sequencing is limited or extra throughput is required.

## Data Availability

All data produced in the present work are contained in the manuscript

## Acknowledgements

ISARIC CCP investigators: Dr Mike Beadsworth, Dr Ingeborg Welters, Dr Lance Turtle, Dr Jane Minton, Karl Ward, Dr Elinor Moore, Dr Elaine Hardy, Dr Mark Nelson, Dr Jane Minton, Karl Ward, Dr David Brealey, Dr Ashley Price, Dr Brian Angus, Dr Graham Cooke and Dr Oliver Koch

