## Supplementary Table 1 for "A Multiplex qPCR to Differentiate Monkeypox Virus Clades"

**Supplementary Table 1**. Primers and probes used in the assay

| **Oligo name** | **Mod5'** | **Sequence** | **Mod3'** | **Concentration** |
| --- | --- | --- | --- | --- |
| Clade I For |  | TCTGTAGGCCGTGTATCAGC |  | 400nM |
| Clade I Rev |  | AAGCTCTGTATGATCTTCAACGT |  | 400nM |
| Clade I Probe | Cy5 | AGGAGTATCGTCGGAACTGTACACCATAGT | BHQ | 200nM |
| Clade Ib For |  | TCCGTTTGATATAGGATGTGGAC |  | 500nM |
| Clade Ib Rev |  | ATATTTGAAACACGGCACTTCG |  | 500nM |
| Clade Ib Probe | Texas Red | CACGTGGGTGGATAATGCGCCTGAATAT | BHQ 2 | 250nM |
| IIb/non-IIb For |  | TCAAAACCAAACGATGATACCTATTCACT |  | 450nM |
| IIb/non-IIb Rev |  | CCAACCTCCACATATTCTGGTTCA |  | 450nM |
| IIb Probe | FAM | TTCAACGATCAAAAAT | MGB | 100nM |
| non-IIb Probe | VIC | AACGGCCAAAAAT | MGB | 100nM |
| Lineage B.1 For |  | TGGAGAAC[G]TCAACGTGTATC |  | 400nM |
| Lineage B.1 Rev |  | GATCACAAG[G]CTGGTACAGA |  | 400nM |
| Lineage B.1 Probe | NED | CATACGATATCTTTGGTATTGA | MGB | 200nM |
| Lineage B.1 PHO blocker | PHO | CATACGATATCTTCGGTATTGA |  | 1mM |
